# Inter-rater reliability of a Chilean-Spanish version of the Post-Intensive Care Unit Presentation Screen (PICUPS): A multicentre study

**DOI:** 10.1101/2025.08.07.25333254

**Authors:** Ignacio Flores-Bustos, Matías Otto-Yáñez, Maria Graciela Salazar-Vega, Jaime Leppe-Zamora, Ana Castro-Avila

## Abstract

**Introduction:** The Post-ICU Presentation Screen (PICUPS) is an assessment tool to identify rehabilitation and referral needs in intensive care unit (ICU) survivors that was recently adapted into Chilean-Spanish. This study aims to determine the inter-rater reliability of different healthcare professionals applying PICUPS (Chilean version) to patients discharged from the ICU.

**Methods:** A multicentre and cross-sectional inter-rater reliability study was conducted among 20 physiotherapist, speech therapists, occupational therapists, and nurses across four ICUs in Chile. Each professional received written instructions, scored two hypothetical clinical cases, and participated in a 1:1 one-hour training session. Five professionals evaluated within 24 hours of ICU discharge patients who received mechanical ventilation during their stay. We estimated reliability using the intraclass correlation coefficient (ICC 1, k) and the average agreement on whether the patient needs a referral.

**Results:** A total of 44 patients were assessed, with a median (P25-P75) age of 61 (43–69) years. The most common admission diagnoses were sepsis (33%) and acute respiratory failure (28%). The PICUPS had excellent inter-rater reliability in 15 of its items (ICC: 0.92–0.98), good reliability in five items (ICC: 0.79–0.87), and moderate reliability in four items (ICC: 0.59–0.74).

**Conclusions:** The PICUPS has adequate inter-rater reliability among professionals working in four ICUs in Chile. These findings are step forward toward the national implementation of this tool for planning the rehabilitation of ICU survivors.

## Introduction

Advancements in intensive care unit (ICU) management have led to increased survival rates among critically ill patients. However, many of these patients experience significant long-term sequelae, collectively referred to as post-intensive care syndrome (PICS)[1]. The reported prevalence of PICS varies across studies, primarily due to differences in assessment methods[2]. A systematic review by Fazzini et al. (2022) estimated that 30% of ICU survivors experienced anxiety at 3 months (95% CI: 19–41%), 24–26% had depression at 3–6 months (95% CI: 17–33%), and their physical function was reduced to 57–63% of predicted values on the 6-minute walk test (95% CI: 45–69%) during the same period [3]. A recent study in Chile reports a similar prevalence, with 36% of ICU survivors experiencing anxiety, 68% physical deterioration, and 37% cognitive impairment at three months [4].

Despite the wide range of health issues affecting ICU survivors, the current literature highlights a global gap in the availability of standardised and validated instruments to assess PICS. This gap is particularly relevant in Spanish-speaking countries, where few tools are accessible in the local language and with proven clinimetric properties. A review by Pant et al. (2023) identified five instruments that evaluated the three core domains of PICS (mental, physical, and cognitive) [5]. However, none of these tools have undergone inter-rater reliability testing, raising concerns about their clinical applicability across different settings. Among these instruments, three (i.e., PICSQ, Healthy Aging Brain Care-Monitor Self Report, and RAIN) have assessed internal validity, one has explored content validity (i.e., Provisional Questionnaire for Long-Term Health-Related Quality of Life and Burden of Disease After Intensive Care), and the last one (i.e., OMIs) has not reported any clinimetric properties. The absence of inter-rater reliability data limits the use of these tools in clinical practice. Adapting and evaluating instruments such as the PICUPS contributes to addressing this shared need by enabling more consistent assessments across diverse clinical contexts.

In Chile, as in many Latin American countries, there are no clinical guidelines or protocols to guide the planning of post-ICU healthcare and rehabilitation [6]. This gap may lead to underestimating patients’ needs, resulting in increased caregiver burden and higher healthcare costs due to delayed rehabilitation and reintegration into the community[7].

The Post-ICU Presentation Screen (PICUPS) was developed during the first wave of the COVID-19 pandemic to identify rehabilitation needs in ICU survivors and to support their reintegration into the community [8]. Until 2024, it was only available in English until Salazar et al. (2023) conducted its translation and cross-cultural adaptation into Chilean-Spanish [9]. Assessing the inter-rater reliability of this tool is crucial for two reasons. Firstly, the user’s manual was developed as part of the cultural adaptation process, but the extent to which it helps standardizing assessment criteria has not yet been tested. Second, since the PICUPS was designed to be used by various healthcare professionals in the ICU, it is essential to determine whether different evaluators apply it consistently. This study aims to determine the inter-rater reliability of the Chilean-Spanish version of the PICUPS among healthcare professionals assessing ICU survivors across four centres.

## Materials and methods

### Study Design

A multicentre and cross-sectional study assessed the inter-rater reliability among 20 healthcare professionals from four intensive care units (ICUs) was conducted from July 9^th^ to November 27^th^, 2024. This study received approval from the Facultad de Medicina Clínica Alemana - Universidad del Desarrollo Research Ethics Committee (No 2023–43) and from the ethics committees of the participating centres. The methodology and results are reported following the Guidelines for Reporting Reliability and Agreement Studies (GRRAS) (S1 Appendix)[10].

### Participants

We invited to participate healthcare professionals who held a degree in nursing, physical therapy, speech therapy, or occupational therapy from a university recognised by the Chilean government and had at least one year of clinical experience in ICUs. All professionals participated voluntarily and provided written informed consent before data collection.

Before assessing patients, participants received written instructions and a one-on-one virtual training session from an ICU physiotherapist that acted as the study coordinator and had received formal training in the use of the PICUPS tool. The training involved scoring the 24 items of the Chilean-Spanish PICUPS based on two hypothetical clinical cases using Google Forms. Subsequently, a virtual meeting was held to discuss the assigned scores and answer queries to ensure standardised scoring across different clinical conditions. In total, participants received 2 hours of training for using the PICUPS. This training was designed to ensure standardisation across clinical disciplines and scenarios and included direct interaction with the trainer to clarify scoring discrepancies.

### Patients

Patients were eligible for inclusion, regardless of admission diagnosis, if they were ≥ 18 years old; received invasive or non-invasive mechanical ventilation for more than 48 hours; and were deemed to be ready for transferring to the high dependency unit. Exclusion criteria included limitation of therapeutic effort and inability to provide voluntary consent due to cognitive or neurological impairments, as determined by a Mini-Mental State Examination (MMSE) score **≤**14 points [11,12] and CAM-ICU positive for delirium [13].

### Instrument

The Post-ICU Presentation Screen (PICUPS) is designed to assess rehabilitation needs in post-ICU patients. It consists of 24 items scored on a 6-point ordinal scale ranging from 0 (most dependent) to 5 (near normal).

The PICUPS basic contains 14 items grouped in four core domains: medical and essential care; breathing and nutrition; physical movement; communication, cognition, and behaviour. The PICUPS plus contains 10 items grouped in three additional domains: upper airway; physical function and activities of daily living; and symptoms interfering with activities.

Each item has a predefined cutoff score indicating the need for referral to physical therapy, speech therapy, psychiatry, and psychology, among others.

The Chilean-Spanish versions of the PICUPS are available as supplementary material (S2 Appendix).

### Data Collection

Each participating centre had a designated physiotherapist responsible for project coordination, recruiting healthcare professionals, managing the informed consent process, and participating in patient evaluations.

Each patient was assessed by five professionals (one physiotherapist, one nurse, one occupational therapist, and one speech therapist). After signing informed consent, the participating professionals received a copy of the Chilean-Spanish PICUPS and the corresponding user’s manual.

The physical therapy coordinator at each centre identified eligible patients and was responsible for obtaining their informed consent. Afterward, the participating professionals independently evaluated each patient within 24 hours, ensuring assessments were not shared among assessors. Responses were recorded via Google Forms.

### Statistical Analysis

Sample size estimation was based on Bonett’s model (2002) [14]. Considering a minimum acceptable reliability of 0.7 (ranging from 0.6 to 0.8), with a 95% confidence interval and five raters per patient the estimated sample size was 57 patients.

Data are presented as absolute and relative frequencies or as median and percentile 25-percentile 75 for the characteristics of the healthcare professionals and patients. Inter-rater reliability was assessed using the intraclass correlation coefficient (ICC 1, k) in a one-way random-effects model to estimate absolute agreement among multiple raters.

The ICC was categorised into distinct levels of reliability: low (< 0.5), moderate (between 0.5 and 0.75), good (between 0.75 and 0.90), and excellent (> 0.90) [15]. Additionally, the median score for each patient and item was used as a reference point to assess agreement regarding referrals to specialists. Based on the score assigned by each evaluator, it was determined whether the assessment led to a specialist referral. The proportion of professionals who made correct referrals was calculated for each patient, and this value was used to compute the average agreement rate. The sign test was employed to determine whether agreements exceeded disagreements.

All statistical analyses were performed using STATA 15.0 SE (StataCorp, College Station, TX, USA).

## Results

This study was conducted from July 9^th^ to November 27^th^, 2024. Each centre had five evaluators who administered the instrument to 44 eligible patients. Five patients were excluded from the statistical analysis because they were assessed by four professionals (Fig 1).

**Figure 1.**
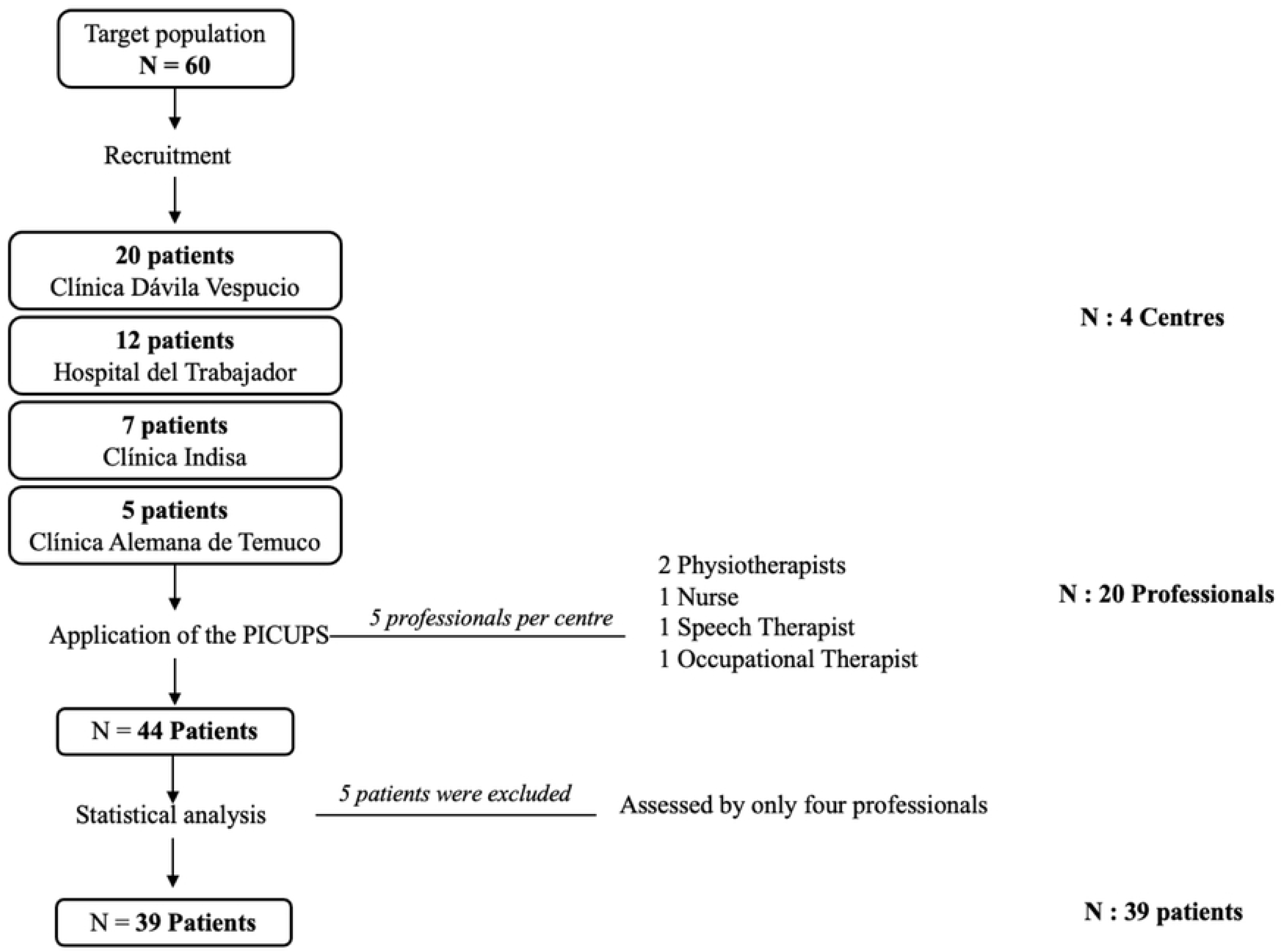
Flowchart of patient enrolment in the study

The group of healthcare professionals was predominantly female (75%) and had a median age of 32 years. Most had between 6 and 10 years of professional experience, with comparable time working in intensive care units. Regarding academic background, 70% held a postgraduate diploma and 30% a master’s degree, indicating a generally high level of training (Table 1).

**Table 1.** Characteristics of the participating professionals.

| Variable | Descriptive statistics |
| --- | --- |
| **Sex (Female/Male)** | 15/5 (75%/25%) |
| **Age (years), median (P<sub>25</sub>-P<sub>75</sub>)** | 32 (30 – 38) |
| **Years of professional experience, median (P<sub>25</sub>-P<sub>75</sub>)** | 7.5 (6 – 10.5) |
| **Years of experience in ICU, median (P<sub>25</sub>-P<sub>75</sub>)** | 6.5 (4.5 – 8.5) |
| **Educational level, n (%)** |  |
| **Diploma** | 14 (70%) |
| **Master's degree** | 6 (30%) |

ICU = Intensive Care Unit.

The patients assessed had a median age of 61 years and were predominantly female (54%). All had undergone prolonged mechanical ventilation, with a median duration of 15 days. The most frequent admission diagnoses were septic shock (33%) and acute respiratory failure (28%), followed by acute brain injury and multiorgan failure, highlighting the severity and complexity of the cohort (Table 2).

**Table 2.** Characteristics of the participating patients.

| Variable | Descriptive statistics |
| --- | --- |
| **Sex M/F** | 18/21 (46%/54%) |
| **Age (years), median (P<sub>25</sub>-P<sub>75</sub>)** | 61 (43-69) |
| **Duration of mechanical ventilation, median P<sub>75</sub>)** | 15 (8-31) |
| **Tracheostomy, n (%)** | 12 (31%) |
| **Admission diagnosis, n (%)** |  |
| **Septic shock** | 13 (33%) |
| **Acute respiratory failure** | 11 (28%) |
| **Acute brain injury** | 5 (13%) |
| **Multiorgan failure** | 3 (8%) |
| **Severe burn** | 2 (5%) |
| **Polytrauma** | 2 (5%) |
| **Neuromuscular disease** | 2 (5%) |
| **Recovered CPR** | 1 (3%) |

TQT = tracheostomy; CPR = cardiopulmonary resuscitation.

Items with excellent inter-rater reliability (ICC > 0.90) included core clinical functions such as respiratory support (Item 3), tracheostomy-related care (Items 4 and 5), swallowing (Item 17), and several mobility and communication items (e.g., Items 6, 8– 11, 16, 18–22). These findings suggest strong consistency across disciplines in assessing these functional domains. Good reliability (ICC 0.75–0.89) was observed in five items, such as medical stability (Item 1), safety and basic needs (Item 2), and care needs (Item 20). These items typically require integrated clinical judgment and may reflect more variability in interpretation across professional backgrounds. Moderate reliability (ICC 0.59–0.74) was observed in items related to emotional and psychosocial domains, such as emotional support needs (Item 14), dyspnoea, fatigue, and pain (Items 15, 23, and 24). These may reflect subjective variations in patient reporting or professional interpretation (Table 3).

**Table 3.**
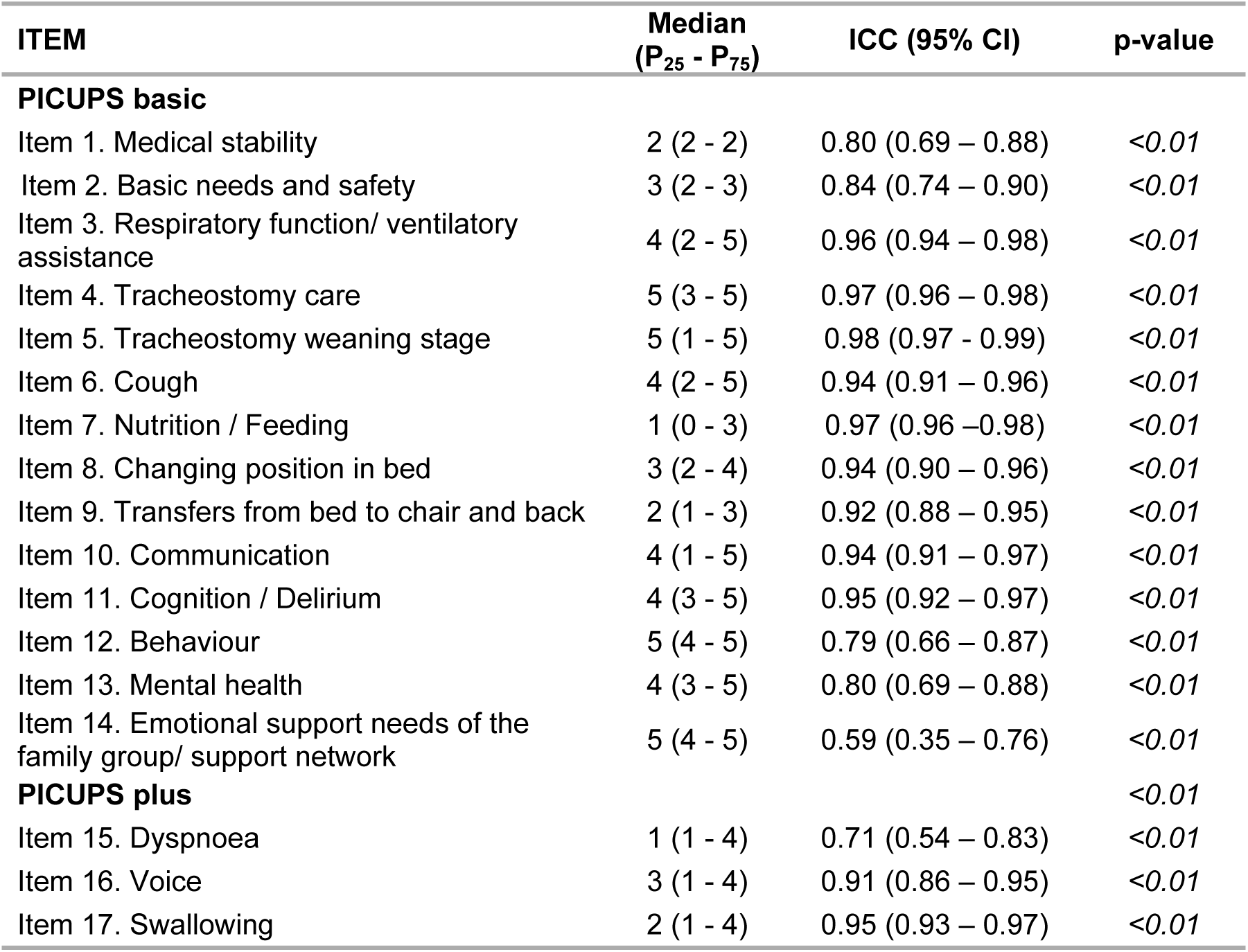
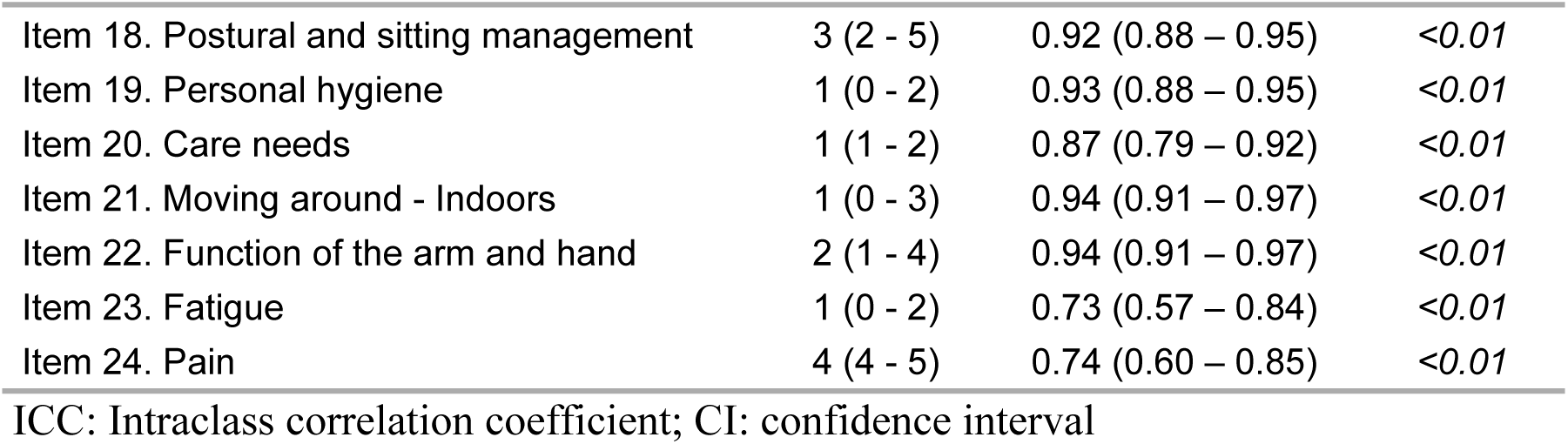
Median scores and ICC per PICUPS item (n = 5 evaluators per centre. Total n= 20).

Agreement among evaluators regarding referral decisions was consistently high across the instrument. All items showed agreement above 80%, with 13 items reaching or exceeding 90%. The highest concordance was observed in tracheostomy care (Item 4) and personal hygiene (Item 19), achieving 99.5% agreement. The lowest agreement was found in the item addressing emotional support needs of the family or support network (Item 14), with 80%, which may reflect greater subjectivity or variability in professional judgment (Table 4).

**Table 4.** Average agreement regarding referral to specialists (n=5 evaluators per centre. Total n= 20).

| ITEM | Average agreement | p-value |
| --- | --- | --- |
| **PICUPS basic** |  |  |
| Item 1. Medical stability | 97.9 | <0.01 |
| Item 2. Basic needs and safety | 99.5 | <0.01 |
| Item 3. Respiratory function/ ventilatory assistance | 93.3 | <0.01 |
| Item 4. Tracheostomy care | 99.5 | <0.01 |
| Item 5. Tracheostomy weaning stage | n.a | n.a |
| Item 6. Cough | 86.7 | <0.01 |
| Item 7. Nutrition / Feeding | 94.4 | <0.01 |
| Item 8. Changing position in bed | 93.8 | <0.01 |
| Item 9. Transfers from bed to chair and back | 97.4 | <0.01 |
| Item 10. Communication | 90.8 | <0.01 |
| Item 11. Cognition / Delirium | 87.2 | <0.01 |
| Item 12. Behaviour | 83.6 | <0.01 |
| Item 13. Mental health | 83.1 | <0.01 |
| Item 14. Emotional support needs of the family group/ support network | 80.0 | <0.01 |
| **PICUPS plus** |  |  |
| Item 15. Dyspnoea | 82.6 | <0.01 |
| Item 16. Voice | 89.2 | <0.01 |
| Item 17. Swallowing | 94.4 | <0.01 |
| Item 18. Postural and sitting management | 89.2 | <0.01 |
| Item 19. Personal hygiene | 99.5 | <0.01 |
| Item 20. Care needs | 99.5 | <0.01 |
| Item 21. Moving around - Indoors | 97.9 | <0.01 |
| Item 22. Function of the arm and hand | 89.7 | <0.01 |
| Item 23. Fatigue | 95.9 | <0.01 |
| Item 24. Pain | 81.0 | <0.01 |

The item “Tracheostomy weaning stage” was excluded from referral analysis, as no referral criterion was defined for this item in the original instrument. Values are presented as percentages (%). n.a = not available.

## Discussion

The Chilean-Spanish PICUPS showed excellent inter-rater reliability in 15 items, moderate in four, and good in five. The level of agreement regarding referral to specialists was above 80% for all instrument items. These reliability levels are similar to those reported for instruments that assess functionality and mobility [16,17].

This is the first study to evaluate the inter-rater reliability of a Spanish-language version of the PICUPS, adapted in Chile. Rather than being limited to a single national context, this version contributes to the broader effort of developing standardised tools for diverse Spanish-speaking populations, particularly in Latin America.

The inter-rater reliability of post-ICU assessment instruments is not always measured. Pant et al. (2023) identified only five tools that assess post-ICU health problems and reported that the developers of these instruments did not test inter-rater reliability [5]. On the other hand, González et al. (2020) reported an ICC of 0.75 among physiotherapists using the FSS-ICU scale to assess functionality in ICU patients [16], while Hiser et al. (2020) found an ICC of 0.98 (95% CI 0.96 - 0.99) among physiotherapists using an instrument that measures patient mobility in the ICU [17]. The lack of information on reliability hinders the comparison and selection of tools suitable for the local context and, therefore, is an area of research that should be further explored.

In hospital settings, ICU survivors are often treated by professionals working in rotating shifts, which may vary depending on staff availability. Therefore, assessment tools must be designed to be used reliably by any available healthcare professional, regardless of discipline or timing. This operational dynamic reinforces the need for instruments that demonstrate strong inter-rater reliability across multidisciplinary teams. However, the multidisciplinary component in inter-rater reliability is rarely explored. Yasumara et al. (2024) determined the inter-rater reliability of the Intensive Care Mobility Scale among 12 physiotherapists and 13 nurses but did not include other professionals working within the ICU [18].

The PICUPS was designed so any healthcare professional could assess the rehabilitation needs of post-ICU patients. However, some instrument items are more specific to a particular profession (e.g., item 16 evaluates voice, or item 22 evaluates function of the arm and hand). Therefore, testing reliability among physiotherapists, nurses, speech therapists, and occupational therapists was considered to increase the instrument’s applicability in clinical practice.

Regarding the timing of detecting post-ICU rehabilitation needs, the Critical Care Society recommends conducting evaluations within 2 to 4 weeks after hospital discharge [19]. However, there is no clear guidance on detecting these needs in the early post-ICU discharge stage. Our study assessed the PICUPS within 24 hours after ICU discharge, so the observed inter-rater reliability reflects what happens in the ward during the early phases of rehabilitation planning. There is limited information on the clinimetric properties of post-ICU assessment tools within these timeframes, as most studies have tested them in patients between 2 and 12 months after ICU discharge.

The application of any assessment tool requires training and/or familiarisation, as well as the standardised criteria for its use. A secondary outcome of this study was the development of a virtual 1:1 training on the instrument for participating professionals, which involved scoring the instrument under different clinical conditions, potentially strengthening the study. The literature provides limited information regarding the duration, content, and optimal format for delivering training on the use of assessment tools. Pant et al. (2023) found that none of the developers of post-ICU scales reported whether the assessors received standardised training before using these scales [5]. Giray et al. (2023) tested the inter-rater reliability of The Chelsea Critical Care Physical Assessment Tool (CPAX) in post-COVID-19 patients but did not report training-related aspects [20]. Only the study by González et al. (2020) reported that the participating received training. Therefore, reporting the characteristics of the training provided in inter-rater reliability studies remains a challenge for future research [16].

The higher inter-rater reliability levels observed for the PICUPS basic compared to the PICUPS plus could be explained by the fact that the assessment was performed immediately after discharge from the ICU. Future studies could focus on evaluating inter-rater reliability at times closer to patient discharge, where it is more challenging for ICU professionals to assess aspects of the PICUPS plus since it evaluates areas less frequently addressed in their clinical practice, such as the level of assistance required for activities of daily living.

This study has some limitations. All four participating ICUs were from private institutions, which may have influenced the characteristics of the professionals and patients. However, since inter-rater reliability is a property of the instrument and the training process rather than the setting, this aspect is unlikely to impact the primary outcome significantly. The sample size calculation was performed using Bonett’s model [14], considering a minimum acceptable reliability of 0.7 (range 0.6 to 0.8), a 95% confidence interval, and five raters per patient, which determined the need to include 57 patients. However, due to time constraints during data collection, achieving the initially estimated sample size was impossible. Nevertheless, our results showed inter-rater reliability levels higher than expected, with 18 items showing an ICC > 0.79 compared to the expected ICC of 0.7. This finding suggests that the lower-than-anticipated variability among raters reduced the potential impact of the smaller sample size. According to Koo and Li (2016), reliability studies should ideally include at least 30 subjects and multiple raters to ensure robust results [21]. However, the authors also acknowledge that when the obtained ICC is higher than expected, it indicates more substantial agreement among raters, which can mitigate the effect of a smaller sample size. Furthermore, Walter et al. (1998) explain that when the ICC is high (≥0.4), the optimal study design often requires fewer observations per subject, indicating that fewer subjects may still yield reliable results [22]. In this context, the high ICC values observed in our study suggest the statistical strength and interpretability of the instrument’s reliability estimates [21,22].

In conclusion, this study found good to excellent inter-rater reliability levels among 20 clinical professionals applying the Chilean-Spanish PICUPS after 1:1 virtual training. This represents a step forward in using the Chilean-Spanish version of the PICUPS to inform the rehabilitation needs of post-intensive care patients in Spanish-speaking settings. While this version was linguistically adapted in Chile, its potential extends to other clinical contexts with similar characteristics across Latin America. Future research should focus on how to implement this tool nationally to determine the rehabilitation needs of this patient population before creating treatment guidelines in the community.

## Data Availability Statement

All relevant data are within the manuscript and its Supporting Information files. Additional data are available from the corresponding author upon reasonable request.

## Notes

### Competing Interest Statement

The authors have declared no competing interest.

### Funding Statement

The author(s) received no specific funding for this work.

### Author Declarations

This study received approval from the Facultad de Medicina Clínica Alemana - Universidad del Desarrollo Research Ethics Committee (No 2023–43) and from the ethics committees of the participating centres. All participants (healthcare professionals and patients) provided oral and written informed consent.

